# Understanding disruptions in cancer care to reduce increased cancer burden

**DOI:** 10.1101/2022.12.26.22283886

**Authors:** Kia L. Davis, Nicole Ackermann, Lisa M. Klesges, Nora Leahy, Walsh-Bailey Callie, Sarah Humble, Bettina Drake, Vetta L. Sanders Thompson

**Author notes:** **Corresponding Author:** Kia Davis, ScD, MPH, Division of Public Health Sciences, Department of Surgery, Washington University School of Medicine, 660 S. Euclid Ave. Campus Box 8100, St. Louis, MO 63110.

## Abstract

**Background:** This study seeks to understand how and for whom COVID-19 disrupted cancer care to understand the potential for cancer health disparities across the cancer prevention and control continuum.

**Methods:** In this cross-sectional study, participants age 30+ residing in an 82-county region in Missouri and Illinois completed an online survey from June-August 2020. Descriptive statistics were calculated for all variables separately and by care disruption status. Logistic regression modeling was conducted to determine the correlates of care disruption.

**Results:** Participants (N=680) reported 21% to 57% of cancer screening or treatment appointments were canceled from March 2020 through the end of 2020. Approximately 34% of residents stated they would need to know if their doctor’s office is taking the appropriate COVID-related safety precautions to return to care. Higher education (OR=1.26, 95%CI:1.11-1.43), identifying as female (OR=1.60, 95%CI:1.12-2.30), experiencing more discrimination in healthcare settings (OR= 1.40, 95%CI:1.13-1.72), and having scheduled a telehealth appointment (OR=1.51, 95%CI:1.07-2.15) were associated with higher odds of care disruption. Factors associated with care disruption were not consistent across races. Higher odds of care disruption for White residents were associated with higher education, female identity, older age, and having scheduled a telehealth appointment, while higher odds of care disruption for Black residents were associated only with higher education.

**Conclusion(s):** This study provides an understanding of the factors associated with cancer care disruption and what patients need to return to care. Results may inform outreach and engagement strategies to reduce delayed cancer screenings and encourage returning to cancer care.

**Funding Support:** This study was supported by the National Cancer Institute’s Administrative Supplements for P30 Cancer Center Support Grants (P30CA091842-18S2 and P30CA091842-19S4). Kia L. Davis, Lisa Klesges, and Bettina Drake were supported by the National Cancer Institute’s P50CA244431 and Kia L. Davis was also supported by the Breast Cancer Research Foundation. Callie Walsh-Bailey was supported by NIMHD T37 MD014218. The content does not necessarily represent the official view of these funding agencies and is solely the responsibility of the authors.

**Availability of data and material:** The dataset generated for the study is not publicly available but is available by request. Interested individuals should contact the corresponding author with a brief description of how the data will be used and proof of IRB approval or exemption. Then a de-identified dataset will be shared.

## Introduction

The COVID-19 pandemic abruptly upended cancer care in many countries including the US. The need to reduce community spread and reserve hospital capacity for the most severe COVID-19 cases led to rescheduling or postponement of cancer care appointments.^1-5^ These control measures significantly decreased cancer-related patient encounters in the early phase of the pandemic, particularly for cancer screenings.^2^ Comparing March to July 2020 with the same period in 2019, there was a substantial decrease in cancer screenings, biopsies, surgeries, office visits, and therapy; the decreases varied by service location and cancer type.^2^ For example, breast cancer screenings decreased by 89.2% and colorectal by 84.5%.^6^ Patients reported delays in receiving cancer care, including follow-up clinic appointments and cancer therapies, such as radiation, infusion therapies, and surgeries.^7,8^

Cancer care delays due to the COVID-19 pandemic are anticipated to lead to increased cancer morbidity and mortality.^9,10^ One study found an association between surgical and screening delays and increased cancer mortality among patients diagnosed with colorectal, lung, and prostate cancer during the pandemic.^4^ Delayed mammography and computed tomography for lung cancer were associated with advanced stage of cancer at diagnosis.^4^ Another study determined delayed surgery for lung cancer was associated with worse survival.^11^ For breast screenings, some evidence suggests that patients were reluctant to return for mammograms after care disruptions.^12^ Thus, cancer care disruptions during COVID-19 could have detrimental future impacts on cancer outcomes and may require changes to public health and clinical strategies across the cancer prevention and control continuum.

It is unclear if patients felt comfortable returning to care in the context of rapidly changing information and guidelines related to COVID-19 and even now that guidelines are more consistent and vaccines are available. There is concern about whether patients will prioritize immediate unmet social needs that might be a result of or exacerbated by COVID-19, such as food insecurity, employment loss, and housing challenges, over disease prevention. Furthermore, people of color, including African Americans, Latinx, and Native communities, as well as those employed in low-wage occupations, are likely to have greater concerns over COVID-19 safety, in addition to the immediate concerns noted above.^13^ Rural communities that already experience limited access to cancer care, have less capacity to manage COVID-19.^14^ Finally, hospitals rapidly increased the use of telehealth to continue cancer care during COVID-19, but older people and those who lived in low-income and rural areas, or were less likely to have commercial insurance were less likely to participate.^15,16^ This combination of factors may exacerbate existing disparities.^13^

This survey study was conducted by NCI-designated Siteman Cancer Center to elucidate: 1) to what extent cancer care appointments (including preventive screenings and treatment) in the bi-state Midwestern catchment area were postponed or canceled, 2) patients’ needs for returning to care, and 3) correlates of care disruption across the catchment area. The cancer burden is significantly greater in this catchment area than the US averages for multiple cancers. Moreover, racial and geographical disparities persist such that African American patients have higher incidence and mortality for lung, colorectal, late-stage breast cancer diagnoses, and prostate cancers compared to White patients. Rural counties also have higher mortality (but not incidence) for melanoma, breast, and prostate cancer compared to urban areas.^17^

Thus, we explore how socio-contextual factors impact cancer health disparities across the continuum of cancer control and prevention during COVID-19 in this bi-state Midwestern catchment area. Race, ethnicity, social class, and gender are social identities that shape many contextual factors related to cancer and COVID outcomes and are considered in this report. We stratify our results by race because of the differential impact of COVID-19 on communities of color and the over-representation of socioeconomic factors such as low-income, low-wage work often experienced by communities of color.^18-20^

## Methods

### Data Source

Data were collected from June through August 2020 as part of Siteman Cancer Center’s Community Outreach and Engagement efforts. The survey focused on understanding cancer prevention and control behaviors throughout the Siteman catchment area. The Siteman catchment area includes 82 counties throughout Missouri (40) and Illinois (42) and is diverse concerning race (21% people of color), geography (15% rural), and healthcare access (29% live in medically underrepresented areas).^21^

### Data Collection

The Washington University in St. Louis, MO Institutional Review Board approved and exempted this study (ID#202006089). We recruited participants through Qualtrics® Online Panels, which emailed potential participants a survey link.^22^ We screened potential participants for the following eligibility criteria: age 30 or older and residing in eastern or southeastern Missouri or central or southern Illinois. Recruitment oversampled for males (35%), people of color (35%) (defined as all races & ethnicities except for non-Hispanic White), and non-metro area residents (20%) (defined as a score of 4 or greater for census-designated rural-urban continuum [RUCC] codes)^23^ to allow for analyses by these groups. The median survey completion time was 20.3 minutes. All participants received an agreed-upon incentive from Qualtrics.

### Measures

#### Outcome variable

Supplemental Table 1 provides detailed information about the measures used in this study. Our outcome of interest, <u>care disruption</u>, was defined as any delay in health or cancer care. Catchment area residents who reported that they decided not to attend an appointment not already canceled due to COVID-19 or they or their doctor/clinic postponed any cancer screening (Pap test, stool blood test, colonoscopy, mammogram, or PSA test) appointment were categorized as experiencing care disruption.

#### Explanatory variables

We included predictor variables that could result in differential access to care due to social stratification: age, race,^24^ ethnicity, gender identity,^25^ sex assigned at birth, sexual orientation, education,^24^ income,^26^ residence in non-metro area, pre-COVID employment, health insurance status, job loss due to COVID-19,^27^ and access to a private vehicle. We also assessed self-report healthcare discrimination using a 7-item scale assessing how many times a participant experienced certain kinds of treatment (overall Cronbach’s alpha = 0.92).^28^ We also controlled for whether they scheduled a telehealth appointment.^29^ All items were adapted from standardized measures, except for sex assigned at birth and access to a private vehicle, which were created by the study team.

We asked if residents participated in a telehealth medical appointment since the COVID-19 pandemic started and whether it was for a general medical appointment or cancer care. While this measure is not directly associated with social stratification, it could be correlated with Internet and other technology access and also predict whether someone was more likely to cancel a scheduled in-person appointment. Finally, we developed a single item to understand what patients who may have experienced care disruption would need *most* to be able to reschedule the appointment. These options included transportation, time to schedule, and knowing: how they would pay for the appointment, if the doctor’s office or clinic was taking appropriate COVID-related safety precautions, if the doctor’s office was still open or scheduling appointments, or that they could bring someone with them; we also included an “other” option with an open-ended response field.

#### Analytic Procedures

Descriptive statistics were obtained for all variables separately and by care disruption status (any care disruption compared to no disruption). Next, logistic regression modeling was conducted to determine the associations with care disruption across the catchment area. For all analyses, “prefer not to answer” responses were recoded as missing. We dropped those who reported that canceling an appointment did not apply to them (n=84) with more males, uninsured people, and those without telehealth appointments reflected in this exclusion. We also used sex at birth and not gender identity in the model due to the near-complete overlap between the two variables and the small sample size for some of the gender-diverse categories (N<6). Additionally, we recoded the job loss variable into the following 3 categories: yes, resident was laid off; no, resident was not laid off; and combined categories of don’t know/not sure/prefer not to answer/not applicable. Finally, we conducted a stratified logistic regression analysis to determine if the associations of care disruption among non-Hispanic Black residents differed when compared to non-Hispanic White residents. Metro/non-metro area was excluded from the stratified non-Hispanic Black and non-Hispanic White models due to a small number of non-Hispanic Black residents in non-metro areas. We do not present other race/ethnicity in the race-stratified models due to the small sample size of participants with non-missing variables for the model in this category (N=71).

## Results

### Sociodemographic and care disruption descriptive information

Unadjusted sociodemographic characteristics of this sample of residents from the Siteman Cancer Center catchment area (n=680) are presented in Table 1. Residents were 46 years old on average. The diverse study sample included 41% respondents of color, and 28% of the respondents live in a non-metro area. The majority of residents identified as female (68%), lived in metro areas (73%), and had a 4-year college or graduate degree (38%). Compared to our catchment area, this sample had a higher proportion of women (68% vs. 51%) and college graduates (38% vs. 30%). We also had a higher proportion of people of color (41% vs. 21%) and residents who lived in rural areas (28% vs. 15%) due to intentional oversampling.

**Table 1.**
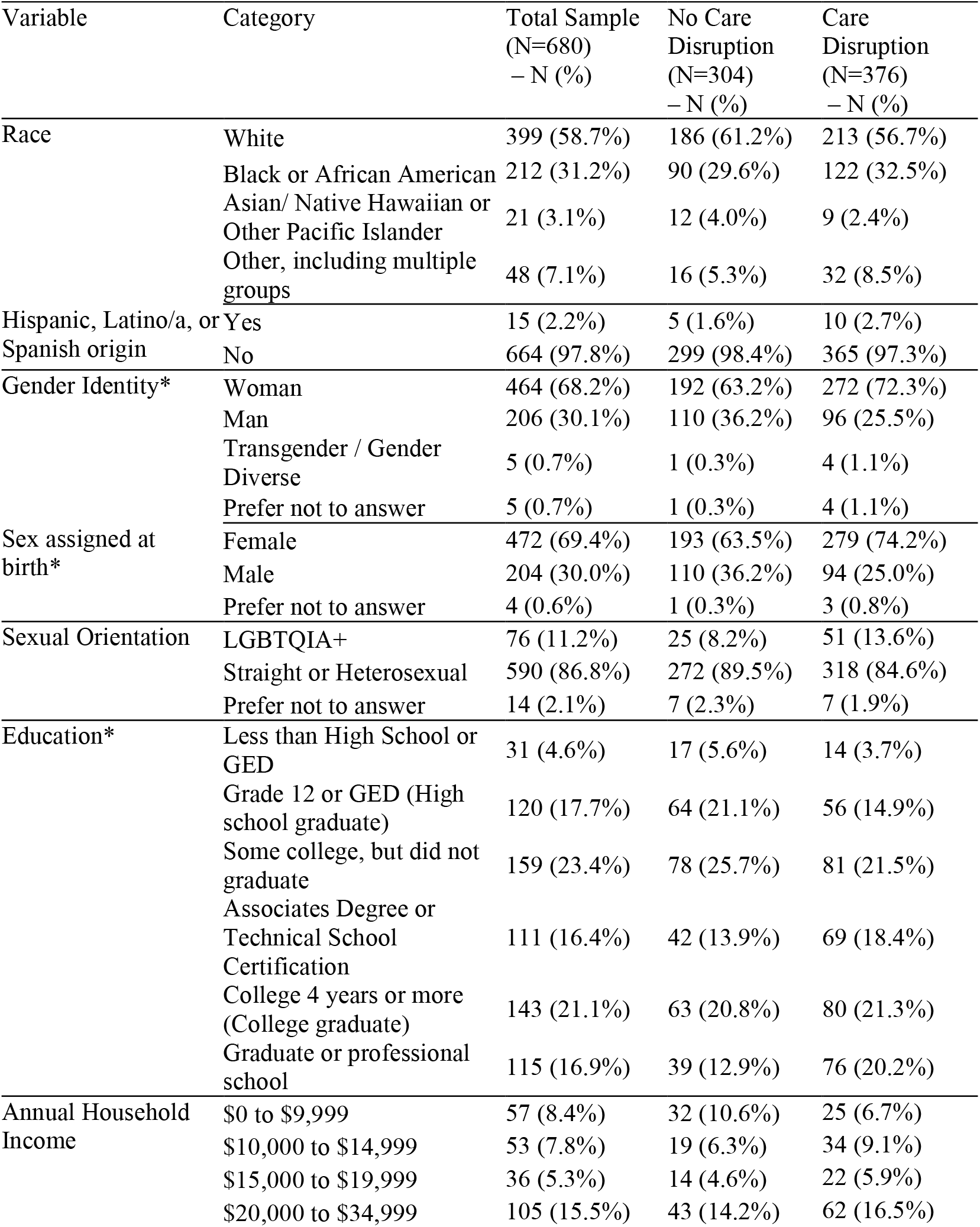
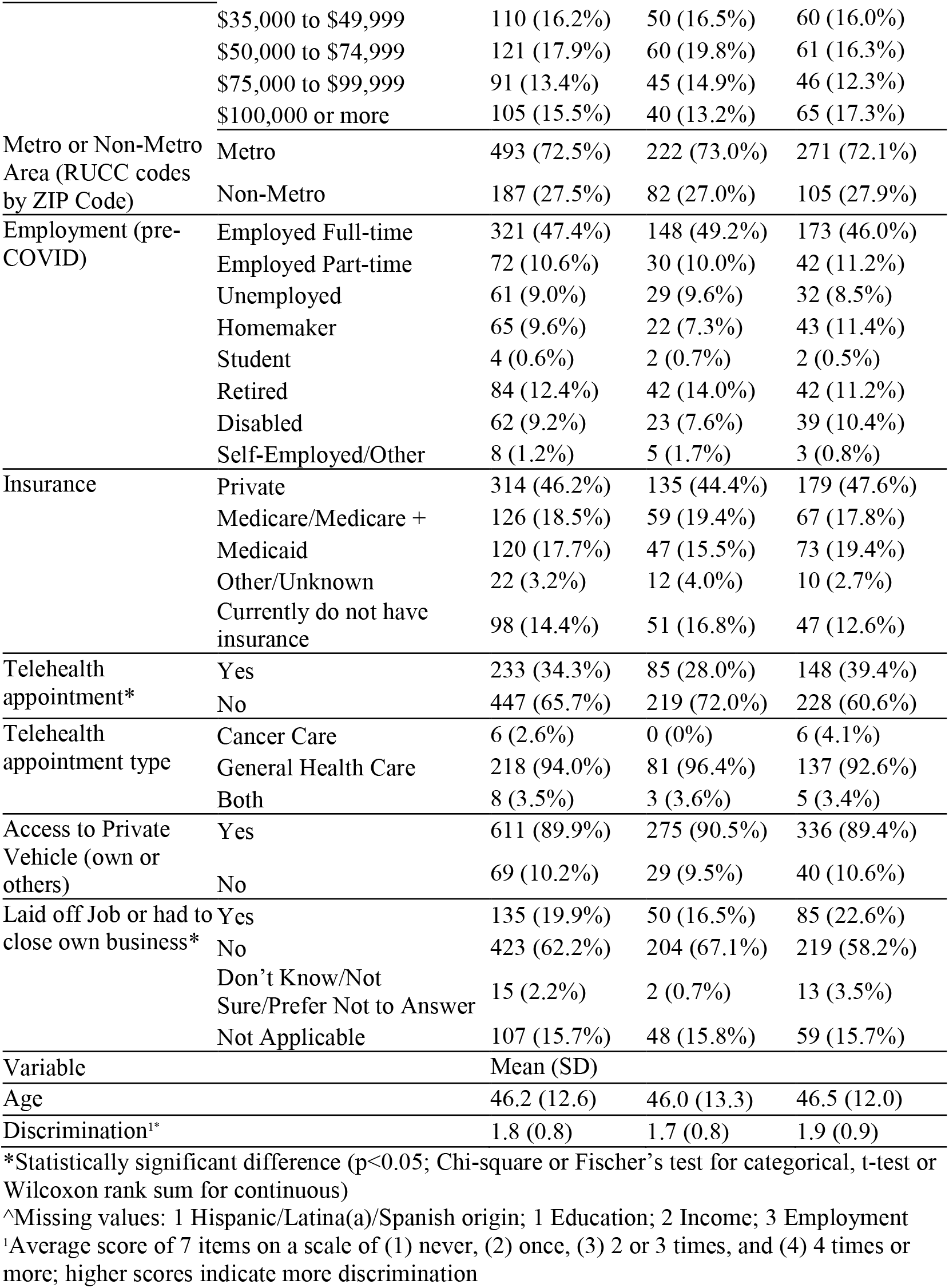
Characteristics of residents across Missouri and Southern Illinois by care disruption status (July-August 2020).

In this sample, approximately 55% of respondents experienced disruption to their scheduled healthcare appointments. Those who experienced care disruption were more likely to be female and have higher levels of educational attainment compared to those who did not experience care disruption.

The number of residents scheduled for a cancer screening appointment, and whose appointment was postponed by the patient or their doctor/clinic is presented in Figure 1. There were 480 possible appointments scheduled between March 2020 through the end of 2020 for either a mammogram, pap test, blood stool test, colonoscopy, or PSA test. Appointment cancelations varied from 21%-57% by screening type. Additionally, in our sample, 25% of residents canceled a scheduled in-person dental appointment, 31% avoided seeking care in a hospital (e.g. labor and delivery, emergency room, etc.), and 46% of residents canceled a scheduled in-person general medical appointment (data not presented).

**Figure 1.**
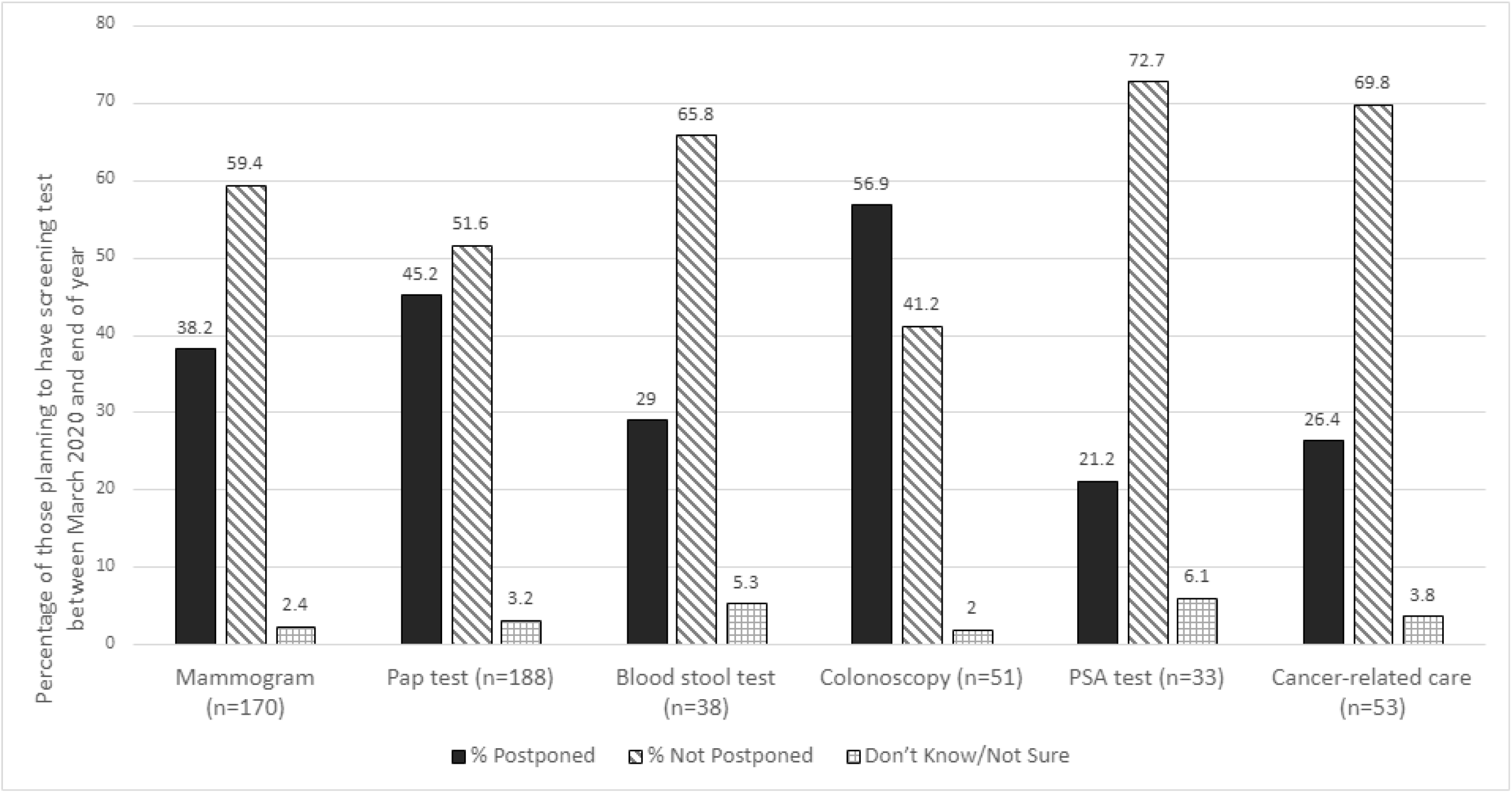
Care Disruption by Cancer Screening/Appointment Type across Missouri and Southern Illinois (July-August 2020) Figure footnotes: N shown is the number who were planning to have a screening test between March 2020 and the end of 2020; For Cancer-related care, this question was asked only of those who self-reported ever being diagnosed as having cancer and the number scheduled for the test is unknown

### Patient needs for rescheduling

In addition, we asked participants who experienced any care disruption what they would need most to reschedule their appointments (n=376). The largest proportion of participants said they would need to know if their doctor’s office or clinic is taking the appropriate COVID-related safety precautions (33.8%), followed by not needing anything (18.1%). Some participants needed to know if their doctor’s office is making appointments for general or routine care (13.3%) or stated they were dealing with other things and not ready to reschedule yet (10.6%). Approximately 8.2% stated they needed to have time to reschedule the appointment. All other needs were reported by less than 5% of respondents.

### Correlates of care disruption

Logistic regression results for the overall and race-specific models are presented in Table 2. In the overall model, higher odds of care disruption were associated with higher educational attainment (OR=1.26, 95% CI: 1.11-1.43), female (OR=1.60, 95% CI: 1.12-2.30), reporting experiencing more discrimination in healthcare settings (OR=1.40, 95% CI: 1.13-1.72), and having scheduled a telehealth appointment (OR=1.51, 95% CI: 1.07-2.15). The correlates of care disruption were not consistent across race. Among Black residents, only higher levels of educational attainment (OR=1.45, 95% CI: 1.13-1.85) were associated with greater odds of care disruption. Whereas, among White residents, higher odds of care disruption were associated with higher levels of educational attainment (OR=1.39, 95% CI: 1.17-1.65), female (OR=1.90, 95% CI: 1.17-3.08), older age (OR=1.02, 95% CI: 1.001-1.04), and having scheduled a telehealth appointment (OR=1.62, 95% CI: 1.01-2.59).

**Table 2.** Odds of any care disruption compared to no care disruption by social factors across Missouri and Southern Illinois (July-August 2020).

| Variable | Overall Sample (N=663) |  | Non-Hispanic Black or African American (N=205) |  | Non-Hispanic White (N=387) |  |
| --- | --- | --- | --- | --- | --- | --- |
|  | Odds Ratio | 95% CI | Odds Ratio | 95% CI | Odds Ratio | 95% CI |
| Race/Ethnicity |  |  |  |  |  |  |
| Non-Hispanic Black or African American | 1.15 | 0.77, 1.72 | -- | -- | -- | -- |
| Other Race/Ethnicity | 1.40 | 0.79, 2.45 | -- | -- | -- | -- |
| Non-Hispanic White (ref) | -- | -- | -- | -- | -- | -- |
| Sex Assigned at Birth<sup>1,3</sup> |  |  |  |  |  |  |
| Female | **1.60** | **1.12, 2.30** | 1.11 | 0.56, 2.19 | **1.90** | **1.17, 3.08** |
| Male (ref) | -- | -- | -- | -- | -- | -- |
| Sexual Orientation |  |  |  |  |  |  |
| LGBTQIA+ | 1.53 | 0.88, 2.65 | 0.68 | 0.27, 1.72 | 1.65 | 0.73, 3.73 |
| Straight or Heterosexual (ref) | -- | -- | -- | -- | -- | -- |
| Area designation (by ZIP code) |  |  |  |  |  |  |
| Non-Metro | 1.23 | 0.82, 1.84 | -- | -- | -- | -- |
| Metro (ref) | -- | -- | -- | -- | -- | -- |
| Telehealth Appointment<sup>1,3</sup> |  |  |  |  |  |  |
| Yes | **1.51** | **1.07, 2.15** | 1.06 | 0.57, 1.99 | **1.62** | **1.01, 2.59** |
| No (ref) | -- | -- | -- | -- | -- | -- |
| Access to Private Vehicle (own or others) |  |  |  |  |  |  |
| Yes | 0.74 | 0.41, 1.33 | 0.79 | 0.33, 1.90 | 0.74 | 0.27, 1.98 |
| No (ref) | -- | -- | -- | -- | -- | -- |
| Health Insurance |  |  |  |  |  |  |
| Medicare/Medicare + | 0.71 | 0.41, 1.24 | 0.88 | 0.31, 2.47 | 0.75 | 0.37, 1.52 |
| Medicaid | 1.02 | 0.59, 1.75 | 0.76 | 0.32, 1.77 | 1.43 | 0.65, 3.15 |
| Other/Unknown | 0.63 | 0.24, 1.66 | 0.75 | 0.15, 3.92 | 0.38 | 0.09, 1.65 |
| Currently do not have insurance | 0.66 | 0.38, 1.13 | 0.58 | 0.22, 1.52 | 0.87 | 0.42, 1.80 |
| Private (ref) | -- | -- | -- | -- | -- | -- |
| Laid off Job or had to close own business |  |  |  |  |  |  |
| Yes | 1.55 | 0.994, 2.41 | 1.55 | 0.75, 3.20 | 1.52 | 0.80, 2.89 |
| No (ref) | -- | -- | -- | -- | -- | -- |
| Don't Know/Not Sure/Prefer Not to Answer/Not Applicable | 1.42 | 0.89, 2.25 | 2.12 | 0.87, 5.18 | 1.06 | 0.59, 1.93 |
| Education<sup>1, 2, 3</sup> | **1.26** | **1.11, 1.43** | **1.45** | **1.13, 1.85** | **1.39** | **1.17, 1.65** |
| Income | 0.99 | 0.89, 1.09 | 0.93 | 0.78, 1.13 | 0.99 | 0.87, 1.13 |
| Discrimination<sup>1</sup> | **1.40** | **1.13, 1.72** | 1.26 | 0.89, 1.78 | 1.29 | 0.96, 1.74 |
| Age<sup>3</sup> | 1.01 | 0.995, 1.03 | 0.99 | 0.96, 1.02 | **1.02** | **1.001, 1.04** |

<sup>1</sup>Statistically significant (p<0.05) overall variable effect – overall model
<sup>2</sup>Statistically significant (p<0.05) overall variable effect – Non-Hispanic Black or African American model
<sup>3</sup>Statistically significant (p<0.05) overall variable effect – Non-Hispanic White model

## Discussion

Using primary data collected from residents across the 82-county Siteman catchment area in Missouri and Illinois, we learned that 21% to 57% of cancer screening or treatment appointments were canceled from March 2020 through the end of 2020. Across all races, residents with higher educational attainment had 1.25 higher odds of care disruption for general or cancer care compared to residents with lower educational attainment; this association remained significant among Black and White residents. Additionally, White residents of older age, assigned female at birth, or having scheduled a telehealth appointment, also had higher odds of care disruption. Finally, knowing their doctor’s office or clinic is taking the appropriate COVID-related safety precautions was the greatest reported need for returning to care (33.8%).

Delays in cancer screening can lead to stage shifts where patients are diagnosed at later stages and thus have a higher risk for cancer morbidity and mortality. Understanding which screenings were impacted and for whom and identifying patient concerns can inform community outreach and engagement efforts. This allows programs to target groups most likely to have delayed screening and draft messaging that can alleviate patient concerns and in turn facilitate a return to care.

Mammograms and Pap tests are an area of increased interest for our catchment area given the high number of women scheduled for screening. Approximately 38% of the 170 women who were scheduled for mammograms had delayed or canceled appointments. Similarly, 45% of the 188 women scheduled for Pap tests had delayed or canceled appointments. Delays in colorectal cancer screening impacted a smaller number of people, but colorectal cancer screening is an important area given the high proportion of cancellations, overall low number of scheduled appointments in general, and high colorectal cancer disparities in the region. Of the 51 people scheduled for a colonoscopy, 57% delayed or canceled appointments, and of the 38 scheduled for a blood stool test, 29% delayed or canceled appointments as well. To help healthcare systems reduce the cancer screening deficit, community outreach and engagement strategies need to address these needs. For example, employing mobile strategies such as the use of mobile mammography and home-based cervical and colorectal cancer screening tests could serve those most impacted.

These data are consistent with prior literature that suggests a reduction in general medical and cancer-related appointments.^2,5,7,30^ This study allows us to understand the magnitude of the impact across Missouri and southern Illinois. Future research exploring whether those with higher educational attainment were more likely to cancel appointments because they were more likely to have better access to scheduling future appointments could further elucidate the extent of educational disparities in healthcare access.

These cross-sectional data cannot infer causality however, many of the correlates of interest (e.g., race, educational attainment) pre-date COVID-19 and the need to consider postponing clinical care. Thus, it is unlikely these results are subject to reverse causation. Also, those excluded due to missing data were more likely to be uninsured. If uninsured persons were also more likely to have postponed appointments, this could potentially bias results about care disruptions by insurance status towards the null and underestimating the impact.

Despite these limitations, this is a significant study that can improve our understanding of COVID-19 impacts on cancer prevention and control and offer specific insights into the region. In our data, those with higher education were more likely to postpone care. This indicates that any trends seen in increasing late-stage diagnosis might occur across socioeconomic categories. Additionally, while Black and White people of higher educational attainment both had increased odds of care disruption, having a scheduled telehealth visit was significantly associated with higher odds of care disruption only for White residents. This suggests that while White people were canceling in-person care, this care may have been substituted with telehealth appointments. Many of these screenings cannot be done virtually, yet this warrants further investigation to understand if care disruption does not always equate to being disconnected from healthcare for some and the subsequent impact on racial disparities in cancer care.

## Data Availability

This was a primary data collection of human subjects data with protected health information. Data is available on request. Interested individuals should contact the corresponding author with a brief description of how the data will be used and proof of IRB approval or exemption. Then a de-identified dataset will be shared.

## Acknowledgments

The authors would like to thank the survey participants for their time, effort, and contribution to the study.

## Appendix

**Supplemental Table 1:**
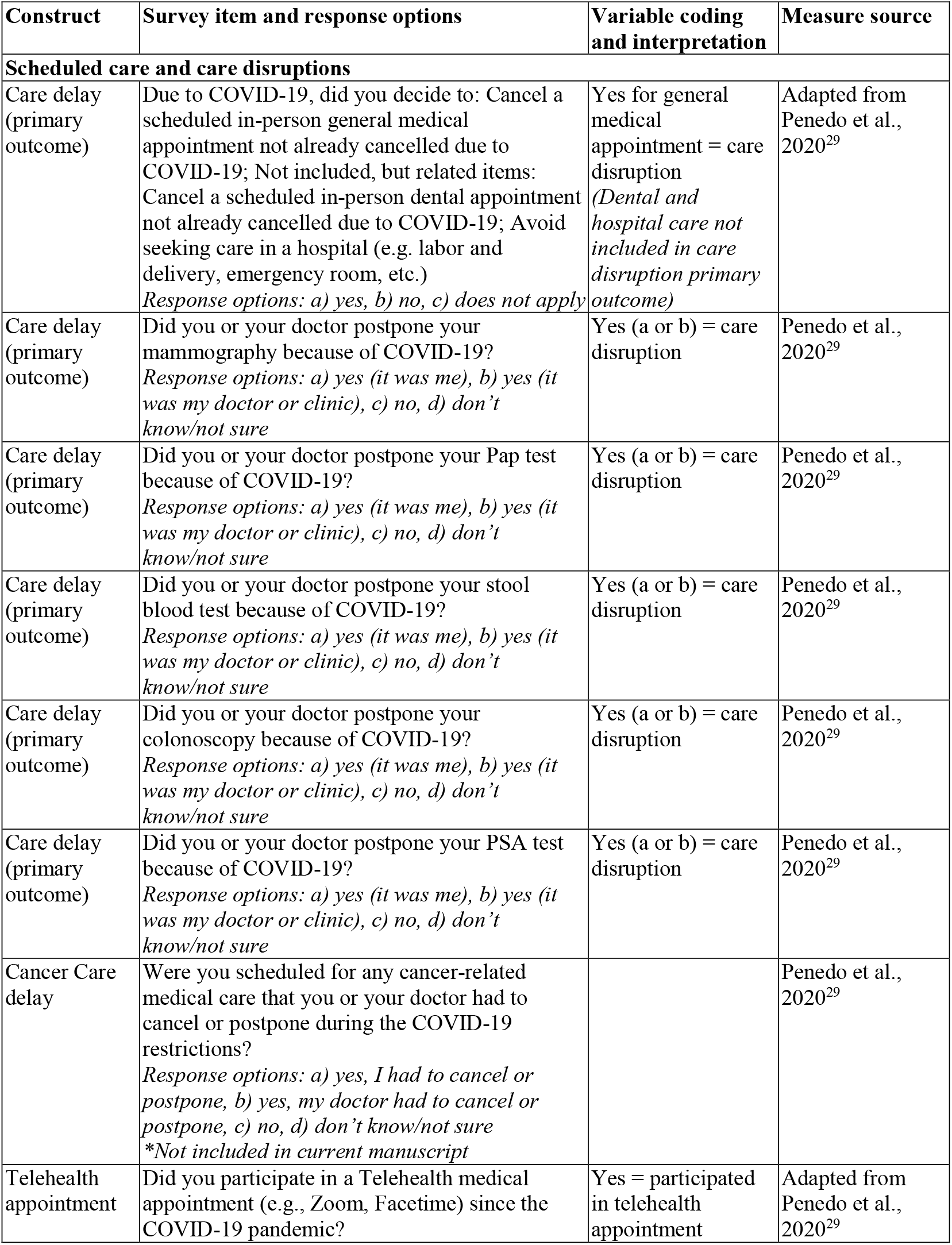
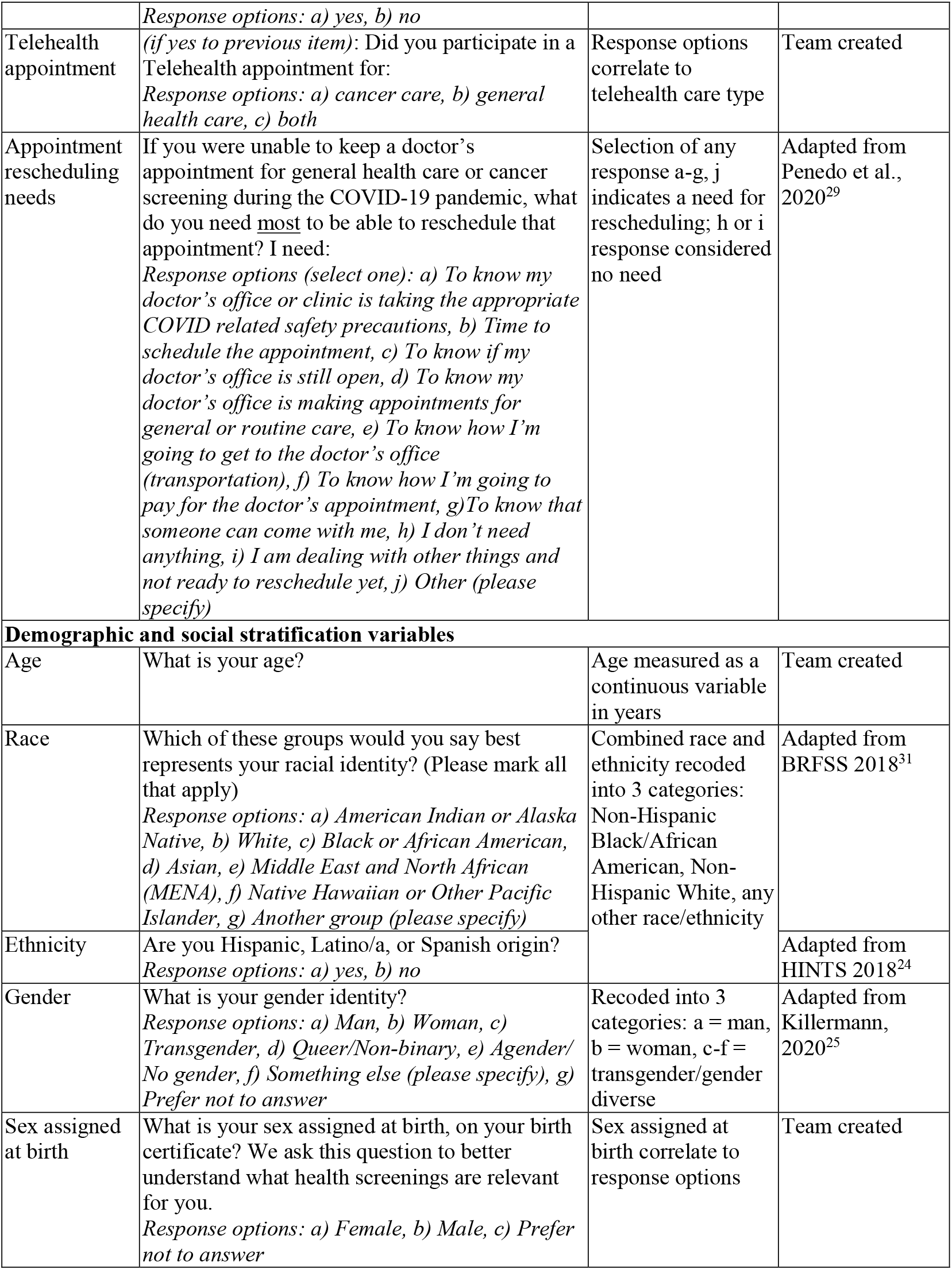
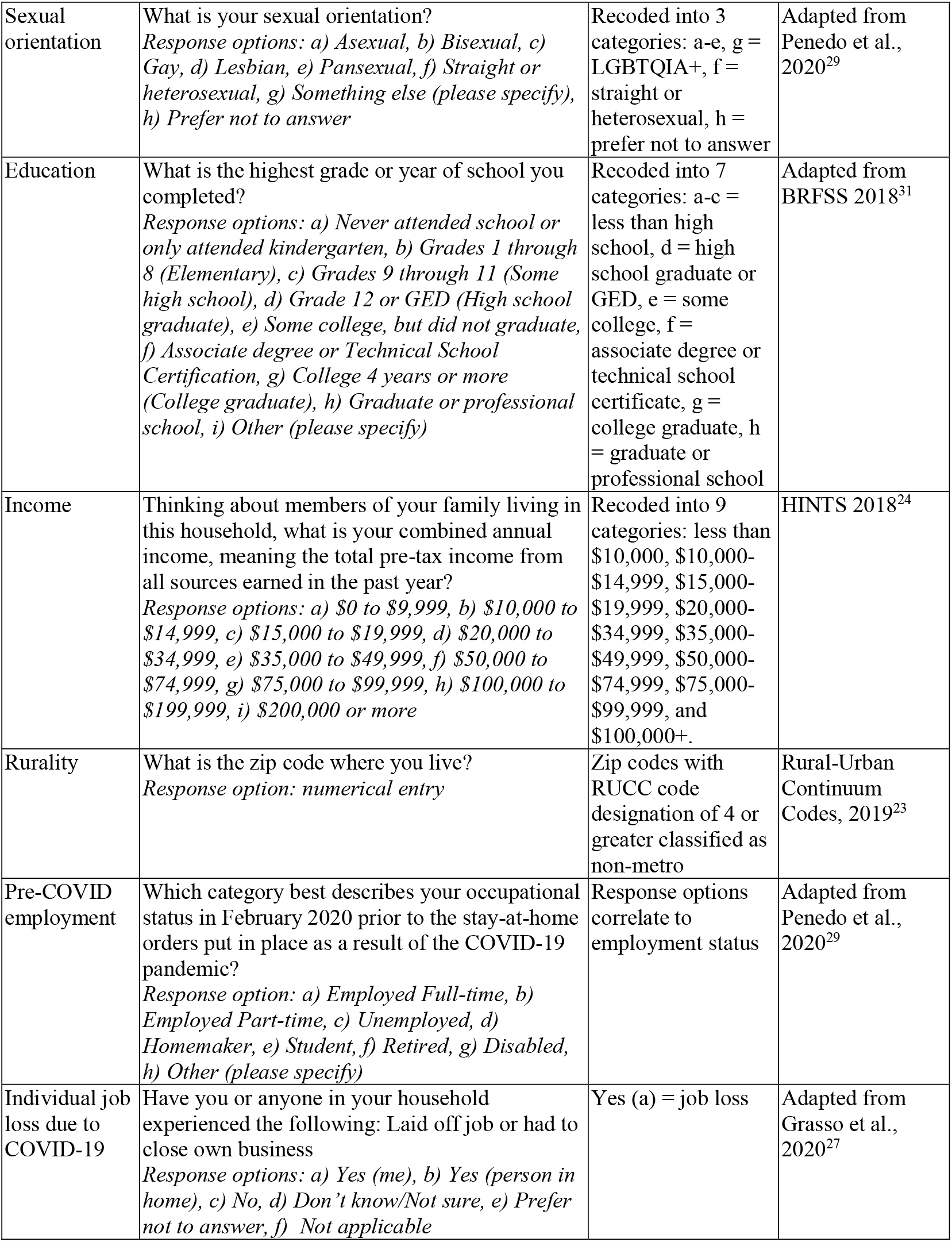
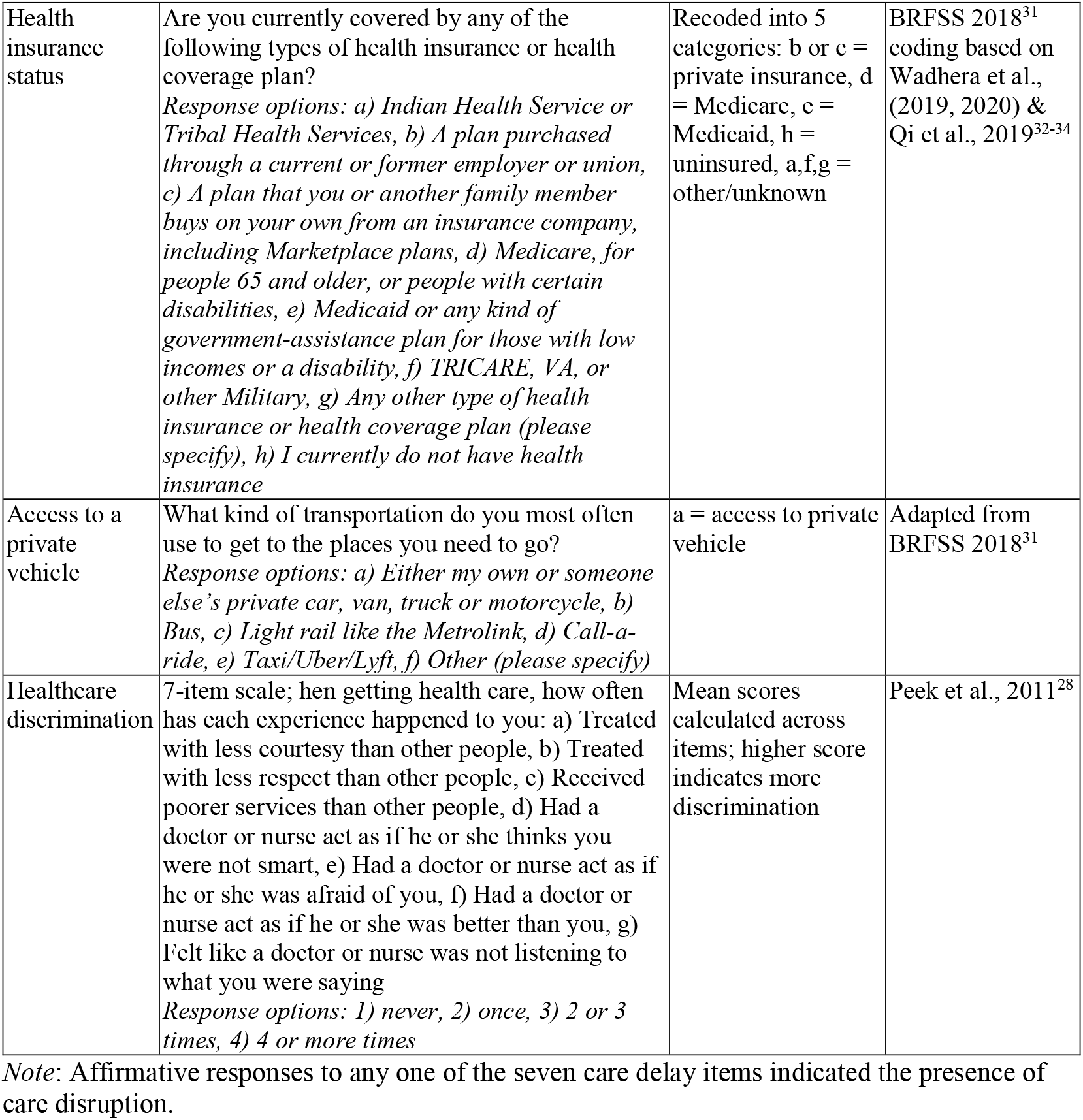
Survey item information

| Construct | Survey item and response options | Variable coding and interpretation | Measure source |
| --- | --- | --- | --- |
| **Scheduled care and care disruptions** |  |  |  |
| Care delay (primary outcome) | Due to COVID-19, did you decide to: Cancel a scheduled in-person general medical appointment not already cancelled due to COVID-19; Not included, but related items: Cancel a scheduled in-person dental appointment not already cancelled due to COVID-19; Avoid seeking care in a hospital (e.g. labor and delivery, emergency room, etc.)<br>*Response options: a) yes, b) no, c) does not apply* | Yes for general medical appointment = care disruption<br>(*Dental and hospital care not included in care disruption primary outcome*) | Adapted from Penedo et al., 2020<sup>29</sup> |
| Care delay (primary outcome) | Did you or your doctor postpone your mammography because of COVID-19?<br>*Response options: a) yes (it was me), b) yes (it was my doctor or clinic), c) no, d) don't know/not sure* | Yes (a or b) = care disruption | Penedo et al., 2020<sup>29</sup> |
| Care delay (primary outcome) | Did you or your doctor postpone your Pap test because of COVID-19?<br>*Response options: a) yes (it was me), b) yes (it was my doctor or clinic), c) no, d) don't know/not sure* | Yes (a or b) = care disruption | Penedo et al., 2020<sup>29</sup> |
| Care delay (primary outcome) | Did you or your doctor postpone your stool blood test because of COVID-19?<br>*Response options: a) yes (it was me), b) yes (it was my doctor or clinic), c) no, d) don't know/not sure* | Yes (a or b) = care disruption | Penedo et al., 2020<sup>29</sup> |
| Care delay (primary outcome) | Did you or your doctor postpone your colonoscopy because of COVID-19?<br>*Response options: a) yes (it was me), b) yes (it was my doctor or clinic), c) no, d) don't know/not sure* | Yes (a or b) = care disruption | Penedo et al., 2020<sup>29</sup> |
| Care delay (primary outcome) | Did you or your doctor postpone your PSA test because of COVID-19?<br>*Response options: a) yes (it was me), b) yes (it was my doctor or clinic), c) no, d) don't know/not sure* | Yes (a or b) = care disruption | Penedo et al., 2020<sup>29</sup> |
| Cancer Care delay | Were you scheduled for any cancer-related medical care that you or your doctor had to cancel or postpone during the COVID-19 restrictions?<br>*Response options: a) yes, I had to cancel or postpone, b) yes, my doctor had to cancel or postpone, c) no, d) don't know/not sure*<br>*\*Not included in current manuscript* |  | Penedo et al., 2020<sup>29</sup> |
| Telehealth appointment | Did you participate in a Telehealth medical appointment (e.g., Zoom, Facetime) since the COVID-19 pandemic? | Yes = participated in telehealth appointment | Adapted from Penedo et al., 2020<sup>29</sup> |
|  | *Response options: a) yes, b) no* |  |  |
| Telehealth appointment | (if yes to previous item): Did you participate in a Telehealth appointment for:<br>*Response options: a) cancer care, b) general health care, c) both* | Response options correlate to telehealth care type | Team created |
| Appointment rescheduling needs | If you were unable to keep a doctor's appointment for general health care or cancer screening during the COVID-19 pandemic, what do you need <u>most</u> to be able to reschedule that appointment? I need:<br>*Response options (select one): a) To know my doctor's office or clinic is taking the appropriate COVID related safety precautions, b) Time to schedule the appointment, c) To know if my doctor's office is still open, d) To know my doctor's office is making appointments for general or routine care, e) To know how I'm going to get to the doctor's office (transportation), f) To know how I'm going to pay for the doctor's appointment, g) To know that someone can come with me, h) I don't need anything, i) I am dealing with other things and not ready to reschedule yet, j) Other (please specify)* | Selection of any response a-g, j indicates a need for rescheduling; h or i response considered no need | Adapted from Penedo et al., 2020<sup>29</sup> |
| **Demographic and social stratification variables** |  |  |  |
| Age | What is your age? | Age measured as a continuous variable in years | Team created |
| Race | Which of these groups would you say best represents your racial identity? (Please mark all that apply)<br>*Response options: a) American Indian or Alaska Native, b) White, c) Black or African American, d) Asian, e) Middle East and North African (MENA), f) Native Hawaiian or Other Pacific Islander, g) Another group (please specify)* | Combined race and ethnicity recoded into 3 categories: Non-Hispanic Black/African American, Non-Hispanic White, any other race/ethnicity | Adapted from BRFSS 2018<sup>31</sup> |
| Ethnicity | Are you Hispanic, Latino/a, or Spanish origin?<br>*Response options: a) yes, b) no* |  | Adapted from HINTS 2018<sup>24</sup> |
| Gender | What is your gender identity?<br>*Response options: a) Man, b) Woman, c) Transgender, d) Queer/Non-binary, e) Agender/No gender, f) Something else (please specify), g) Prefer not to answer* | Recoded into 3 categories: a = man, b = woman, c-f = transgender/gender diverse | Adapted from Killermann, 2020<sup>25</sup> |
| Sex assigned at birth | What is your sex assigned at birth, on your birth certificate? We ask this question to better understand what health screenings are relevant for you.<br>*Response options: a) Female, b) Male, c) Prefer not to answer* | Sex assigned at birth correlate to response options | Team created |
| Sexual orientation | What is your sexual orientation?<br>*Response options: a) Asexual, b) Bisexual, c) Gay, d) Lesbian, e) Pansexual, f) Straight or heterosexual, g) Something else (please specify), h) Prefer not to answer* | Recoded into 3 categories: a-e, g = LGBTQIA+, f = straight or heterosexual, h = prefer not to answer | Adapted from Penedo et al., 2020<sup>29</sup> |
| Education | What is the highest grade or year of school you completed?<br>*Response options: a) Never attended school or only attended kindergarten, b) Grades 1 through 8 (Elementary), c) Grades 9 through 11 (Some high school), d) Grade 12 or GED (High school graduate), e) Some college, but did not graduate, f) Associate degree or Technical School Certification, g) College 4 years or more (College graduate), h) Graduate or professional school, i) Other (please specify)* | Recoded into 7 categories: a-c = less than high school, d = high school graduate or GED, e = some college, f = associate degree or technical school certificate, g = college graduate, h = graduate or professional school | Adapted from BRFSS 2018<sup>31</sup> |
| Income | Thinking about members of your family living in this household, what is your combined annual income, meaning the total pre-tax income from all sources earned in the past year?<br>*Response options: a) \$0 to \$9,999, b) \$10,000 to \$14,999, c) \$15,000 to \$19,999, d) \$20,000 to \$34,999, e) \$35,000 to \$49,999, f) \$50,000 to \$74,999, g) \$75,000 to \$99,999, h) \$100,000 to \$199,999, i) \$200,000 or more* | Recoded into 9 categories: less than \$10,000, \$10,000-\$14,999, \$15,000-\$19,999, \$20,000-\$34,999, \$35,000-\$49,999, \$50,000-\$74,999, \$75,000-\$99,999, and \$100,000+. | HINTS 2018<sup>24</sup> |
| Rurality | What is the zip code where you live?<br>*Response option: numerical entry* | Zip codes with RUCC code designation of 4 or greater classified as non-metro | Rural-Urban Continuum Codes, 2019<sup>23</sup> |
| Pre-COVID employment | Which category best describes your occupational status in February 2020 prior to the stay-at-home orders put in place as a result of the COVID-19 pandemic?<br>*Response option: a) Employed Full-time, b) Employed Part-time, c) Unemployed, d) Homemaker, e) Student, f) Retired, g) Disabled, h) Other (please specify)* | Response options correlate to employment status | Adapted from Penedo et al., 2020<sup>29</sup> |
| Individual job loss due to COVID-19 | Have you or anyone in your household experienced the following: Laid off job or had to close own business<br>*Response options: a) Yes (me), b) Yes (person in home), c) No, d) Don't know/Not sure, e) Prefer not to answer, f) Not applicable* | Yes (a) = job loss | Adapted from Grasso et al., 2020<sup>27</sup> |
| Health insurance status | Are you currently covered by any of the following types of health insurance or health coverage plan?<br>*Response options: a) Indian Health Service or Tribal Health Services, b) A plan purchased through a current or former employer or union, c) A plan that you or another family member buys on your own from an insurance company, including Marketplace plans, d) Medicare, for people 65 and older, or people with certain disabilities, e) Medicaid or any kind of government-assistance plan for those with low incomes or a disability, f) TRICARE, VA, or other Military, g) Any other type of health insurance or health coverage plan (please specify), h) I currently do not have health insurance* | Recoded into 5 categories: b or c = private insurance, d = Medicare, e = Medicaid, h = uninsured, a,f,g = other/unknown | BRFSS 2018<sup>31</sup> coding based on Wadhera et al., (2019, 2020) & Qi et al., 2019<sup>32-34</sup> |
| Access to a private vehicle | What kind of transportation do you most often use to get to the places you need to go?<br>*Response options: a) Either my own or someone else's private car, van, truck or motorcycle, b) Bus, c) Light rail like the Metrolink, d) Call-a-ride, e) Taxi/Uber/Lyft, f) Other (please specify)* | a = access to private vehicle | Adapted from BRFSS 2018<sup>31</sup> |
| Healthcare discrimination | 7-item scale; when getting health care, how often has each experience happened to you: a) Treated with less courtesy than other people, b) Treated with less respect than other people, c) Received poorer services than other people, d) Had a doctor or nurse act as if he or she thinks you were not smart, e) Had a doctor or nurse act as if he or she was afraid of you, f) Had a doctor or nurse act as if he or she was better than you, g) Felt like a doctor or nurse was not listening to what you were saying<br>*Response options: 1) never, 2) once, 3) 2 or 3 times, 4) 4 or more times* | Mean scores calculated across items; higher score indicates more discrimination | Peek et al., 2011<sup>28</sup> |

*Note:* Affirmative responses to any one of the seven care delay items indicated the presence of care disruption.

